## Supplemental for "Linking multiple serological assays to infer dengue virus infections from paired samples using mixture models"

*Dengue virus nested reverse-transcriptase polymerase chain reaction (Nested RT-PCR)*

The DENV nested RT-PCR was performed following the method previously described by Klungthong et al. 2015. The method comprised two sequential RT-PCR rounds: the 1^st^ round RT-PCR and the subsequent 2^nd^ round nested RT-PCR. For the 1^st^ round RT-PCR, a pair of universal DENV forward and reverse primers were used. This step involved a single RT-PCR procedure, initiated with a RT step at 42°C for 60 minutes, followed by RT-PCR amplification for 35 cycles with the thermocycling conditions of denaturation at 94°C for 30 seconds, annealing at 55°C for 1 minute, and extension at 72°C for 2 minutes. Following the completion of the 1^st^ round RT-PCR, the RT-PCR products were specifically diluted and utilized as the template for the subsequent 2^nd^ round nested RT-PCR. The nested RT-PCR assay employed a mixture of five primers, including the universal forward primer used in the 1st round RT-PCR and four DENV type-specific reverse primers. The 2^nd^ round nested RT-PCR was performed using 25 cycles with the same thermocycling conditions as the 1^st^ round RT-PCR step. After the nested RT-PCR amplification, the RT-PCR products were subjected to agarose gel electrophoresis for analysis. The presence of specific DNA bands on the gel enabled the identification of DENV specimens containing types 1, 2, 3, or 4. Specifically, the detection of a DNA band of 482, 119, 290, and 392 base pairs (bp) indicates DENV-1, DENV-2, DENV-3, and DENV-4, respectively. These DNA bands were compared to the amplified DNA from positive controls, which represent the DENV genome. The composition of the RT-PCR buffer mixture and the primers were described previously ^17^.

*Hemagglutination inhibition assay (HAI)*

HAI was carried out using goose erythrocytes as previously described ^41^. In short, sucrose-acetone extracted DENV1 (Hawaii), DENV-2 (NGC), DENV-3 (H87), DENV-4 (H241), and JEV (JaGAr01) antigens from suckling mouse brain have been used as hemagglutinating antigens. Test sera, positive and negative controls were serial 2-fold diluted starting from 1:10 and extending to 1:20,480 and then 250 µL of each dilution was transferred into a v-bottom 96-well plate (Thermo Scientific™, US). Then, an equal volume of antigen (8-16 HA units) was added and incubated at 4^o^C overnight before adding 500 µL of goose red blood cells. After incubation at room temperature for 2 hours, the hemagglutination reaction was observed. A non-inhibitory titer of 1:10 is annotated as <10.

*Anti-dengue/JE IgM/IgG enzyme immunoassay*

Anti-DENV/JEV IgM and IgG capture EIA was used in this study and performed in duplicate wells of 96-well flat-bottom microplate. Briefly, microplates were coated with 100 μL/well of 1:1,600 dilution of goat anti-human IgM or IgG (KPL, Gaithersburg, MD) in 0.018 M carbonate buffer (pH 9.0). After overnight incubation at 4°C, the plates were washed with phosphate buffered saline (PBS) (pH 7.4) containing 0.5% Tween 20 (PBS-T). Next, 50 μL/well of 1:100 dilution of test serum, negative control (NC), weak positive control (WPC), and strong positive control (SPC) in PBS were added and incubated overnight at 4°C. After washing with PBS-T, 50 μL/well of sucrose acetone extracted suckling mouse brain DENV (pooled DENV antigen: DENV-1 [Hawaii], DENV-2 [NGC], DENV-3 [H87], and DENV-4 [H241]) and JEV (JaGAr01) antigens were added into DENV (IgM/IgG) and JEV (IgM/IgG) plates, respectively. After incubation for 2 h at room temperature, 30 μL/well of human anti-flavivirus IgG–horseradish peroxidase conjugated was added and incubated for 1 hour at 37°C. After washing with PBS-T, 100 μL/well of TMB substrate (KPL, Gaithersburg, MD) was added and incubated for 10–30 min. The reaction was stopped by adding 50 μL/well of 0.2 M sulfuric acid. The absorbance (optical density [OD]) was measured at a wavelength of 450 nm (SoftMax Pro Software, Molecular Devices, San Jose, CA). A valid assay should provide OD values at < 0.100, 0.400–0.600, and > 0.600 for NC, WPC, and SPC, respectively. EIA units of tested serum are equal to 100 × [(ODTest − ODNC)/(ODWPC − ODNC)];

*Classification as primary or post-primary DENV infection*

Primary DENV infection was defined by a 4-fold rise and HAI titers at or below 1,280. Post-primary DENV infection was defined by a 4-fold rise and HAI titers at or higher than 2,560. Recent DENV infections were defined by HAI titers at or higher than 2,560 at the acute sampling event. For EIA interpretations units of IgM ≥ 40 were used as a positive cut-off value. Evidence of DENV infection was classified by a ratio of DENV IgM/JEV IgM ≥ 1.0, and JEV infection when the ratio was < 1.0. Primary DENV infection was interpreted when the ratio of DENV IgM/DENV IgG was ≥ 1.8, and post-primary DENV infection was considered when the ratio was < 1.8.

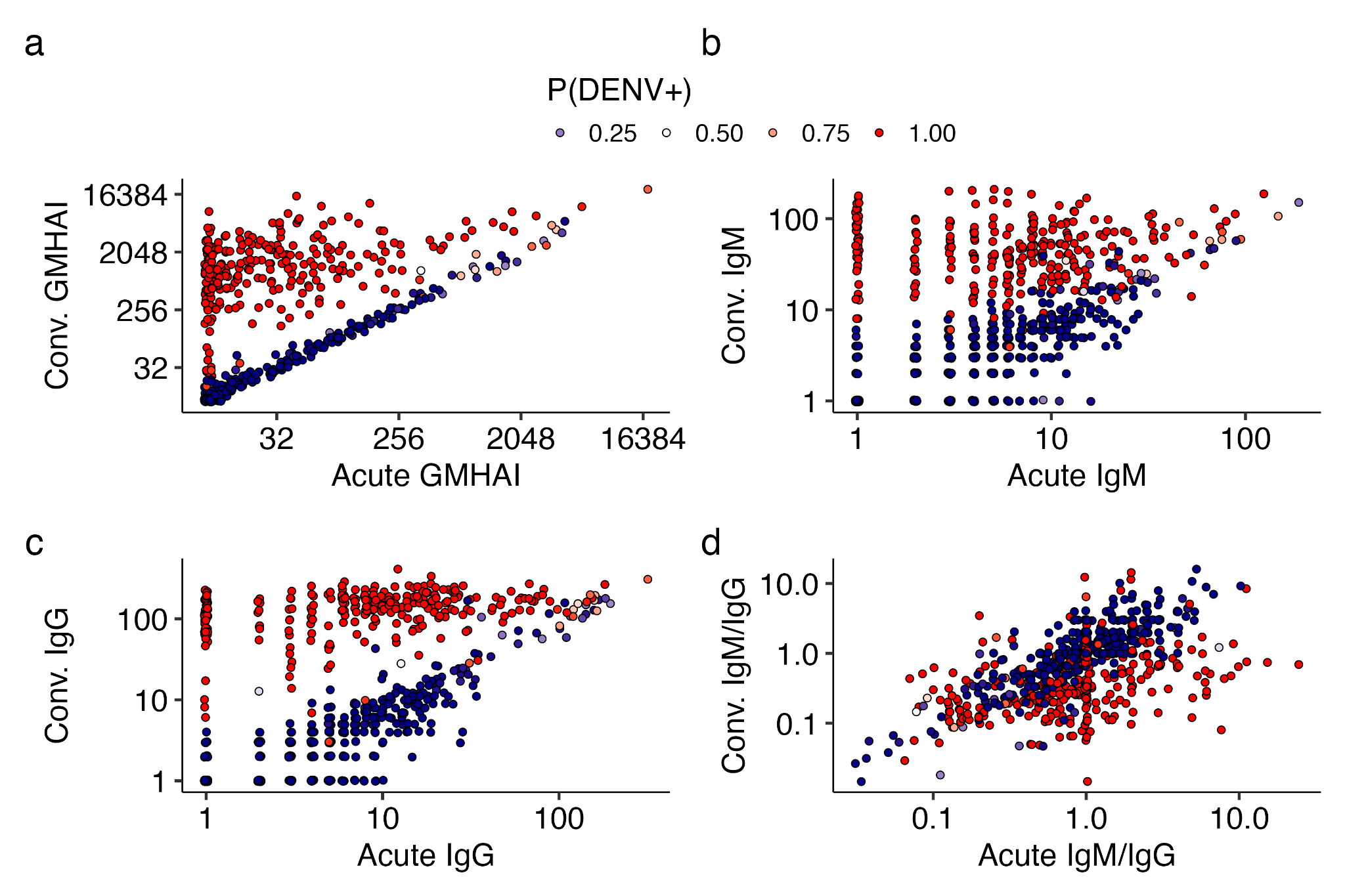

**Figure S1.** Acute and convalescent data for serological assays, colored by probability of infection found during model testing for a model that accounts for time since symptom onset. a) Geometric mean titers of a haemagglutination inhibition assay (GM HAI) for all four serotypes of dengue virus. b) Immunoglobulin G (IgG) c) Immunoglobulin M (IgM) d) Ratio of IgM to IgG at both acute and convalescent sera samples.

**Table S1**. Parameters for forward simulation using serosim^37^. Unless otherwise noted all parameters are drawn from log-normal distributions.

| **Parameter name** | **Definition** | **Simulated mean** | **Simulated standard deviation** |
| --- | --- | --- | --- |
| boost_long | Titer increase post infection  (long term post infection) | GM HAI: 4  IgG: 5  IgM: 5 | GM HAI: 1  IgG: 30  IgM: 30 |
| boost_short | Titer increase post infection (quickly post infection) | GM HAI: 0.75  IgG: 80  IgM: 50 | GM HAI: 0.1  IgG: 30  IgM: 70 |
| wane_long | Rate of titer decrease linearly with time (days) | GM HAI: 0.0001  IgG: 0.005  IgM: 0.005 | GM HAI: 0.001  IgG: 0.005  IgM: 0.005 |
| wane_short | Rate of titer decrease linearly with time (days) | GM HAI: 0.005  IgG: 0.02  IgM: 0.0075 | GM HAI: 0.001  IgG: 0.05  IgM: 0.05 |
| biomarker_prot_midpoint | The biomarker quantity at which you are 50% protected from infection | GM HAI: 8  IgG: NA  IgM: NA | NA |
| biomarker_prot_width | Determines the shape of the curve | GM HAI: .8  IgG: NA  IgM: NA | NA |
| upper_bound | Max titer for biomarker | GM HAI: 20480  IgG: 500  IgM: 500 | NA |
| lower_bound | Min titer for biomarker | GM HAI: 10  IgG: 0  IgM: 0 | NA |
| obs_sd (drawn from Gaussian) | Observation standard deviation – lowest noise | GM HAI: 0.05  IgG: 1.5  IgM: 1.5 | NA |
|  | Observation standard deviation – additional noise | GM HAI: 0.15  IgG: 2.5  IgM: 2.5 | NA |
|  | Observation standard deviation – highest noise | GM HAI: 0.3  IgG: 5  IgM: 5 | NA |
| biomarker_ceiling_threshold | The maximum biomarker level an individual can have before their boost is limited in size | GM HAI: 10  IgG: 80  IgM: 75 | NA |
| biomarker_ceiling_gradient | (1-A)/B; Where A is the proportion of the full boost received at or above the biomarker_ceiling_threshold (B) | GM HAI: 0.15  IgG: 0.01  IgM: 0.01 | NA |
| max_events | Number of infection events possible | 4 | NA |

**Table S2.** Model comparisons for simulated data with differing amounts of observational noise. Percentages around no noise, low noise, and high noise represent the amount of noise relative to the baseline set by the realistic noise scenario. The realistic noise scenario had observational noise set to 0.15, 0.4, and 0.4 for HAI, IgG, and IgM respectively. For models with convergence issues an NA is presented.

|  | **Infection definition** | | | | | | | | | | | |
| --- | --- | --- | --- | --- | --- | --- | --- | --- | --- | --- | --- | --- |
|  | **Lowest noise** | | | | **Additional noise** | | | | **Highest noise** | | | |
| **Model** | Sens. | Spec. | F1 | AUC ROC | Sens. | Spec. | F1 | AUC ROC | Sens. | Spec. | F1 | AUC ROC |
| IgM | 55.7% | 99.7% | 71.3% | 93.6% | NA | NA | NA | NA | NA | NA | NA | NA |
| IgG | 67.6% | **100%** | 80.7% | 93.1% | 63.1% | **100%** | 77.4% | 92.0% | 59.7% | **100%** | 74.7% | 94.9% |
| GM HAI | 95.5% | 99.1% | 96.8% | 98.4% | 90.3% | 97.5% | 92.7% | 97.0% | 80.7% | 94.4% | 84.5% | 90.2% |
| GM HAI + IgM | 97.7% | 99.7% | 98.6% | 99.9% | 94.9% | 99.4% | 96.8% | 99.7% | 86.4% | 95.7% | 88.9% | 97.5% |
| GM HAI + IgG | 97.7% | 97.8% | 96.9% | 99.8% | 95.5% | 99.4% | 97.1% | 99.7% | 90.3% | 97.8% | 93.0% | 98.2% |
| IgM + IgG | 76.1% | **100%** | 86.5% | 98.1% | 73.9% | 99.7% | 84.7% | 97.3% | 68.8% | 99.7% | 81.2% | 94.7% |
| GM HAI + IgG + IgM | **98.3%** | 98.8% | **98.0%** | **99.9%** | **96.6%** | **100%** | **98.3%** | **99.9%** | **91.5%** | 98.1% | **93.9%** | **99.0%** |

**Table S3.** Distribution of paired serological data that fall into probability buckets when using Model 9 (incorporating paired IgG, IgM, GM HAI, and acute GM HAI serological data) and the number of serological assays (RT-PCR, HAI, and EIA) that interpret the individual as having an infection.

|  | **Number of positive interpretations** | | | |
| --- | --- | --- | --- | --- |
| **P(DENV+)** | **0** | **1** | **2** | **3** |
| **[0,0.25)** | 360 | 20 | 2 | 0 |
| **[0.25,.5)** | 0 | 0 | 0 | 0 |
| **[0.5,.75)** | 1 | 2 | 2 | 2 |
| **[.75,1]** | 2 | 17 | 63 | 206 |

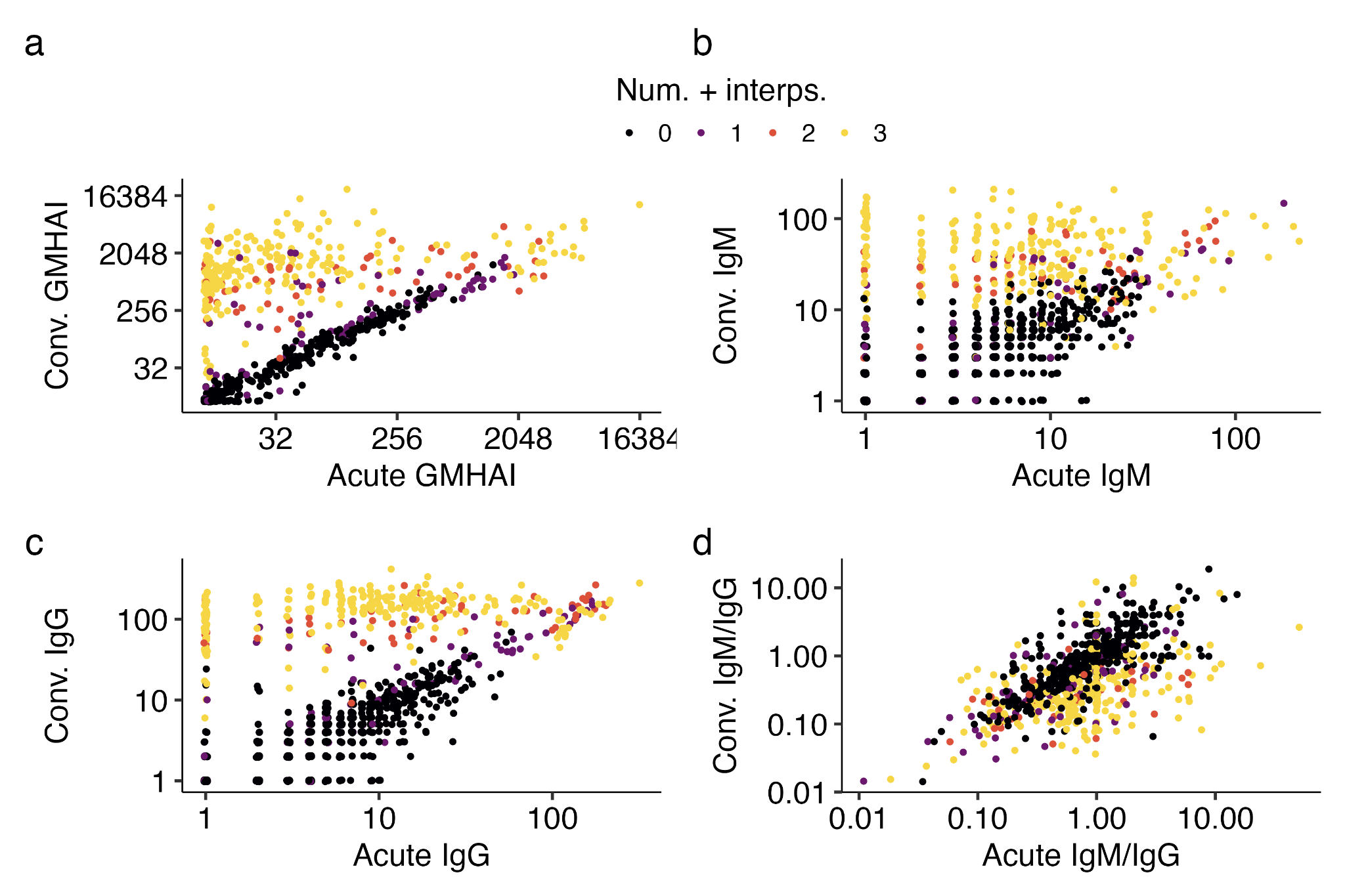

**Figure S2.** Acute and convalescent samples from cohort study data for serological assays, colored by the number of serological assays (RT-PCR, HAI, and EIA) that interpret the individual as having an infection. a) Geometric mean titers of a haemagglutination inhibition assay (GM HAI) for all four serotypes of dengue virus. b) Immunoglobulin G (IgG) c) Immunoglobulin M (IgM) d) Ratio of IgM to IgG at both acute and convalescent serum samples.

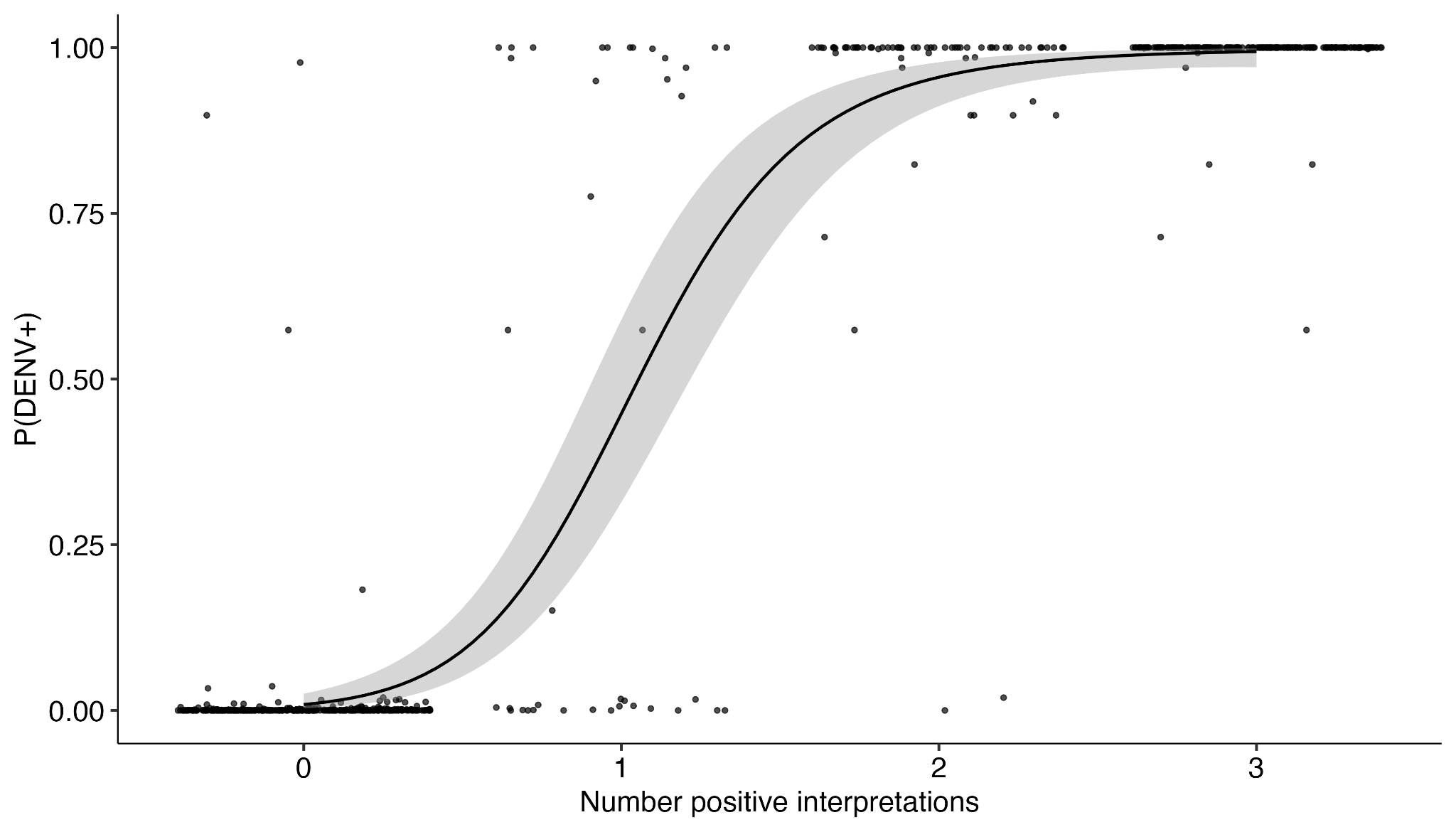

**Figure S3.** Relationship between estimated probability of a dengue virus infection (DENV) found during model testing for Model 9 (incorporating paired IgG, IgM, GM HAI, and acute GM HAI serological data) and the number of serological assays (RT-PCR, HAI, and EIA) that interpret the individual as having an infection.

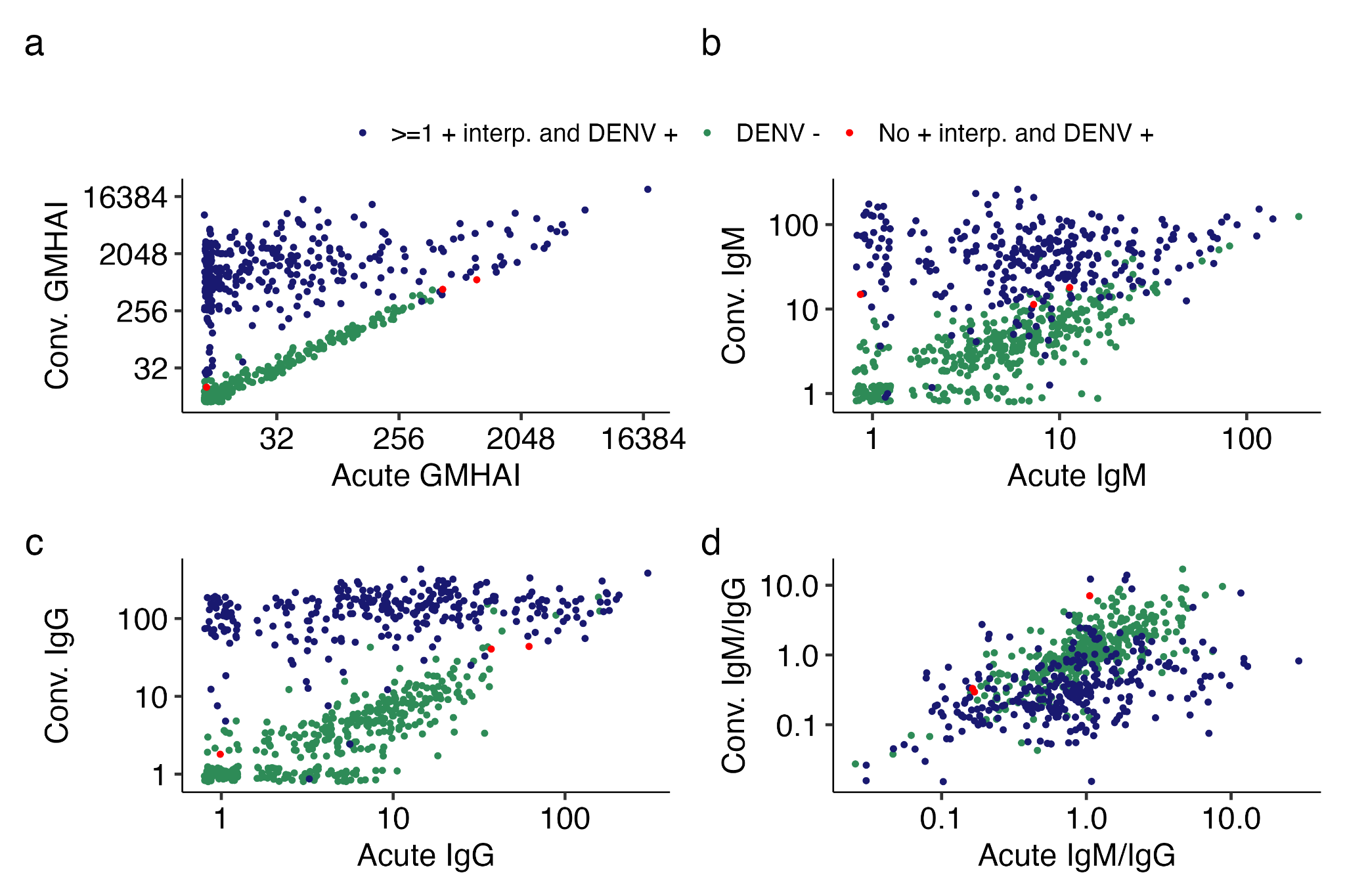

**Figure S4.** Acute and convalescent samples from cohort study data for serological assays, colored by whether they are predicted as infections using Model 9. Green points represent no predicted infection while samples classified as an infection are split into two groups, those with no positive serological interpretations (RT-PCR, HAI, and EIA) in red while those with at least one positive serological interpretation are blue. a) Geometric mean titers of a haemagglutination inhibition assay (GM HAI) for all four serotypes of dengue virus. b) Immunoglobulin G (IgG) c) Immunoglobulin M (IgM) d) Ratio of IgM to IgG at both acute and convalescent serum samples.

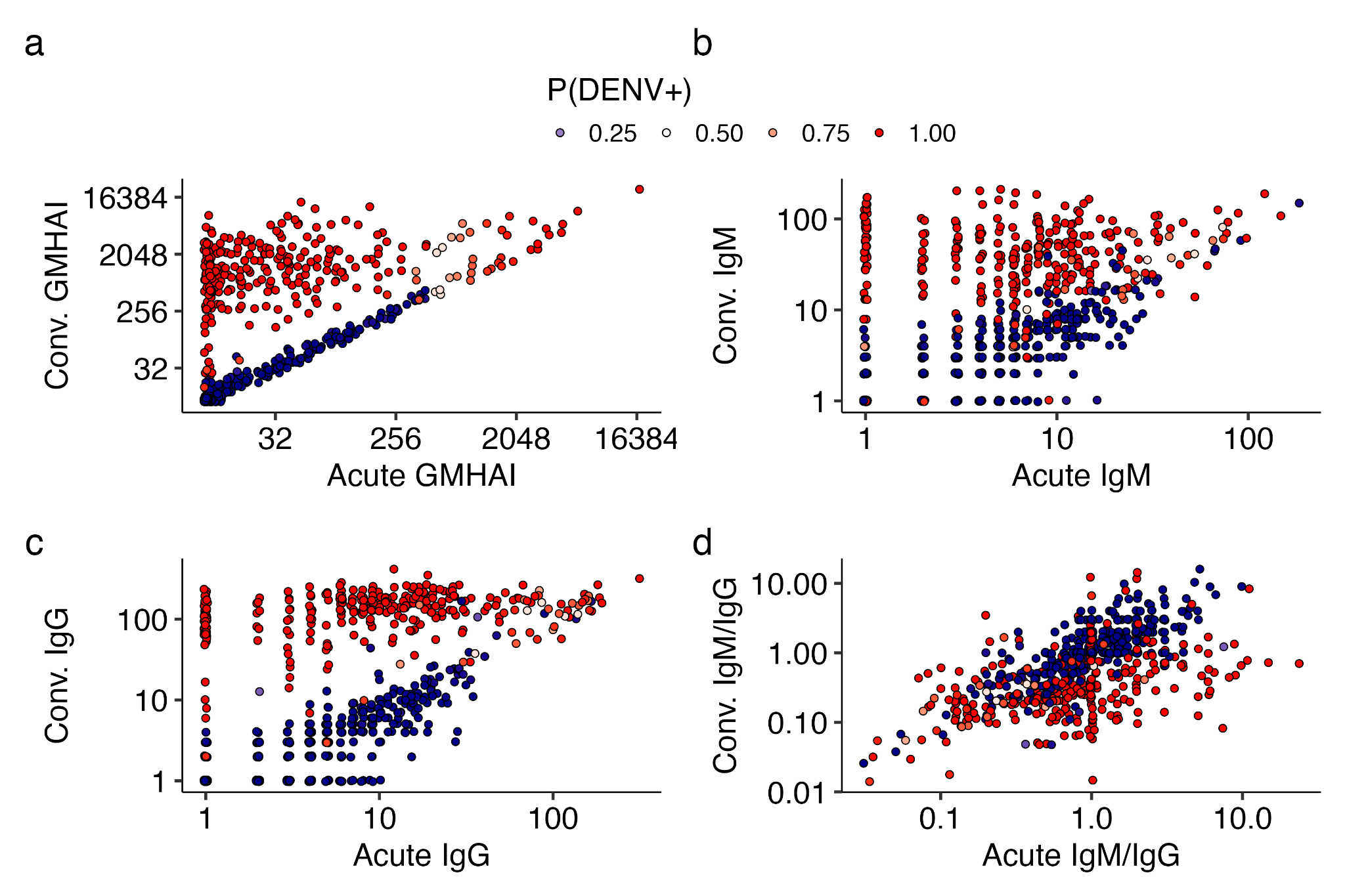

**Figure S5.** Acute and convalescent data for serological assays, colored by probability of infection found during model testing for Model 9. a) Geometric mean titers of a haemagglutination inhibition assay (GM HAI) for all four serotypes of dengue virus. b) Immunoglobulin G (IgG) c) Immunoglobulin M (IgM) d) Ratio of IgM to IgG at both acute and convalescent sera samples.

**Table S4.** Comparison of accuracy metrics (sensitivity, specificity, and AUC ROC) of each model defined by what subset of data was used for training and testing. For models with convergence issues an NA is presented.

| **Model** | | **Infection definition** | | | | | | | |
| --- | --- | --- | --- | --- | --- | --- | --- | --- | --- |
|  |  | RT-PCR+ vs. RT-PCR - | | | | RT-PCR, HAI, or EIA + vs.  RT-PCR, HAI, and EIA - | | | |
| # | Data | Sens. | Spec. | F1 score | AUC ROC | Sens. | Spec. | F1 score | AUC ROC |
| – | HAI clinical interp. | 98.3% | 82.5% | 83.7% | – | *94.8%* | *100%* | *97.4%* | *–* |
| – | EIA clinical interp. | 90.1% | 85.6% | 81.8% | – | *84.5%* | *100%* | *91.6%* | *–* |
| 10 | Acute IgG | 6.0% | **90.9%** | 9.7% | 53.5% | 17.3% | **99.7%** | 29.4% | 63.9% |
| 11 | Conv. IgG | 96.1% | 55.3% | 66.4% | **92.6%** | 91.8% | 64.7% | 78.7% | **95.5%** |
| 12 | Acute IgM | 13.4% | 86.0% | 18.7% | 51.8% | 21.8% | 93.0% | 33.6% | 61.4% |
| 13 | Conv. IgM | 97.0% | 57.6% | **67.9%** | 91.3% | 91.5% | 66.8% | **79.4%** | 93.8% |
| 14 | Acute  GM HAI | 56.3% | 28.8% | 37.0% | 42.8% | 65.2% | 32.5% | 53.3% | 56.3% |
| 15 | Conv.  GM HAI | **98.3%** | 29.4% | 56.8% | 90.9% | **94.5%** | 33.2% | 69.3% | 94.1% |

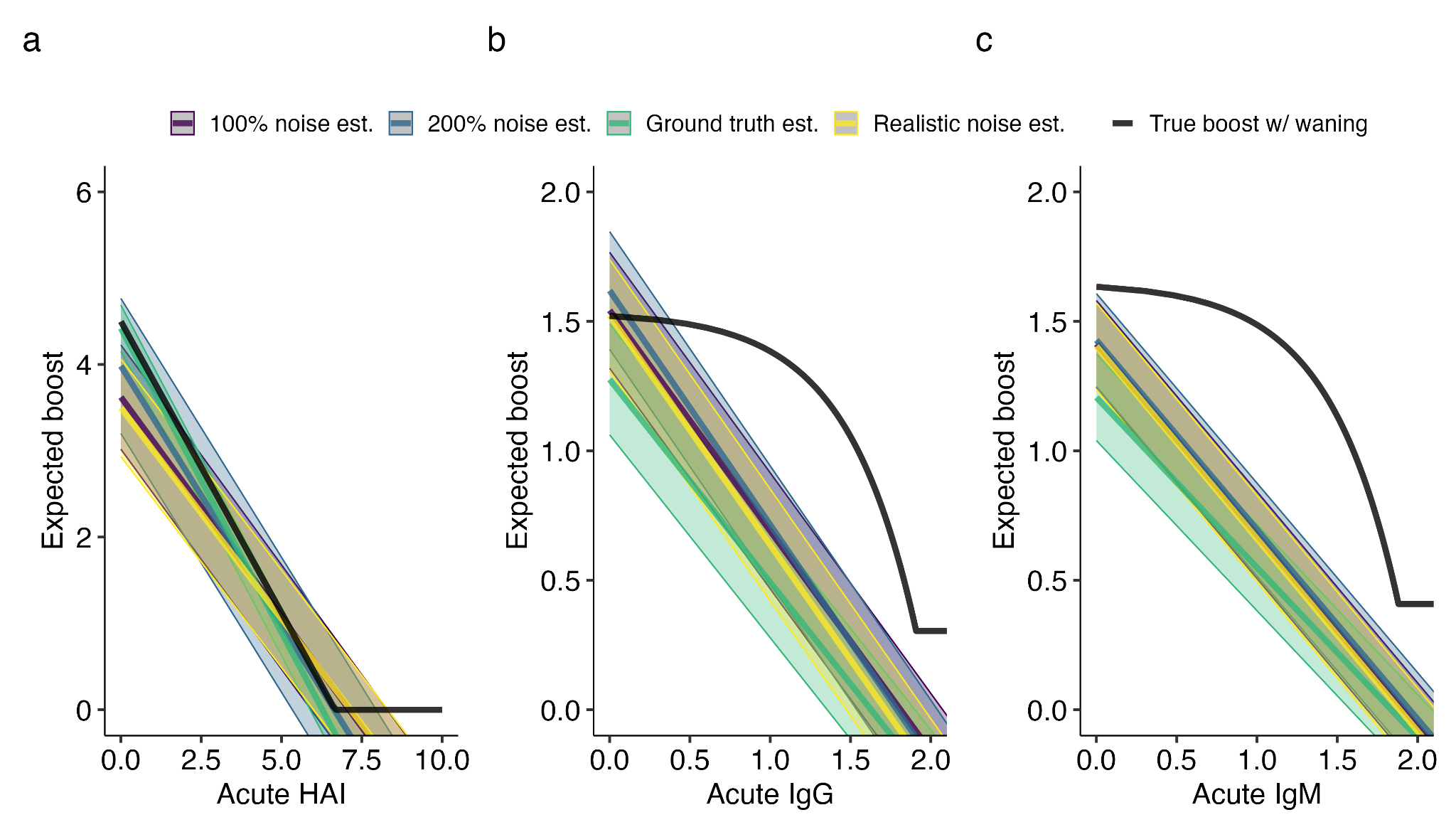

**Figure S6.** Comparison between simulation based estimates for boosts in GM HAI, IgG, and IgM that is experienced after an infection event as a function of acute GM HAI, IgG, and IgM titers individuals under the varying levels of observational noise. The black solid lines represent the defined functional relationship between acute and convalescent titers after the boost and subsequent waning of titers between the samples.

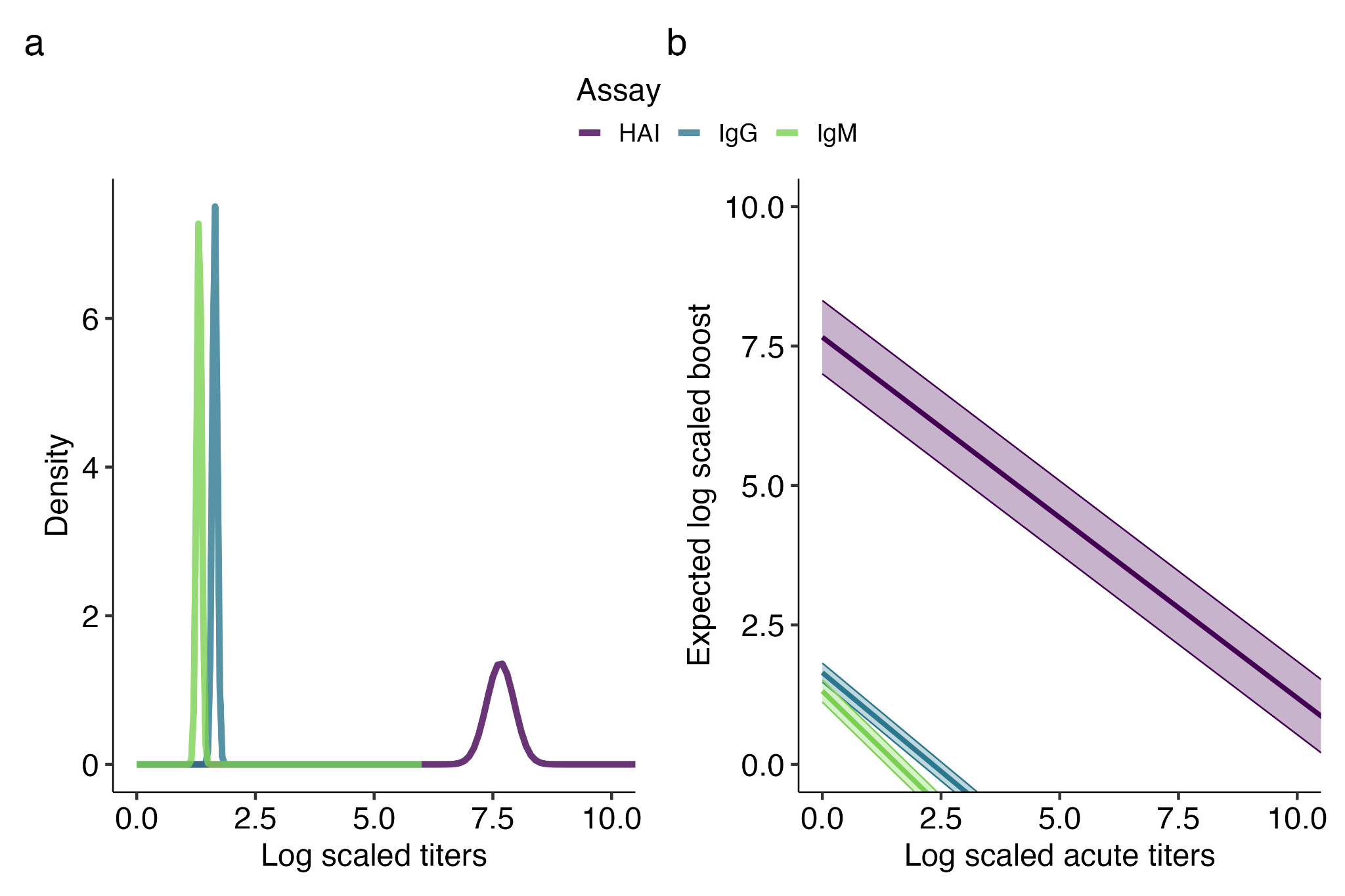

**Figure S7.** (A) Estimated titer boost a fully susceptible individual receives upon a primary infection for each assay. (B) Estimated relationship between titer boost and acute titer for each assay.
